## Supporting information File 1 for "PREOPERATIVE CLINICAL PRACTICE GUIDELINES FOR ELECTIVE SURGERY. METHODOLOGICAL AND QUALITY APPRAISAL STUDY"

**Supplementary Material File 2**

**Table. Characteristics of Clinical Practice Guidelines, stratified by recommendation status* (n=20)**

| **Characteristic** | **Recommended CPG**  **n=14**  **No (%)** | **Recommended with modifications CPG**  **n=6**  **No (%)** |
| --- | --- | --- |
| Main topic  Preoperative fasting (n=6)  Cardiac assessment for non-cardiac surgery (n=11)  Use of routine preoperative tests (n=3) | 4 (67)  8 (73)  2 (67) | 2 (33)  3 (27)  1 (33) |
| Year of publication  2010 (n=1)  2011 (n=5)  2014 (n=3)  2016 (n=3)  2017 (n=4)  2018 (n=1)  2019 (n=1)  2020 (n=1)  2021 (n=1) | 1 (100)  3 (60)  2 (67)  1 (33)  4 (100)  1 (100)  0 (0)  1 (100)  1 (100) | 0 (0)  2 (40)  1 (33)  2 (67)  0 (0)  0 (0)  1 (100)  0 (0)  0 (0) |
| Number of authors  ≤ 5 (n=1)  6 – 10 (n=5)  11 – 20 (n=6)  ≥ 20 (n=8) | 0 (0)  4 (80)  4 (67)  6 (75) | 1 (100)  1 (20)  2 (33)  2 (25) |
| Type of institution  Governmental institution (n=2)  Specialty society or consortium (n=16)  University or academic institution (n=2) | 2 (100)  10 (63)  2 (100) | 0 (0)  6 (37)  0 (0) |
| Region  United States and Canada (n=5)  Europe (n=7)  Latin America (n=5)  Other regions (n=3) | 5 (100)  6 (86)  2 (40)  1 (33) | 0 (0)  1 (14)  3 (60)  2 (67) |
| Guideline version  First version (n=12)  Revision or updated version (n=8) | 9 (75)  5 (63) | 3 (25)  3 (37) |
| Reported funding  No (n=10)  Yes (n=10) | 4 (40)  10 (100) | 6 (60)  0 (0) |
| Method for guideline development  Systematic review (n=19)  Not mentioned (n=1) | 14 (73)  0 (0) | 5 (27)  1 (100) |
| Recommendation methods  Not mentioned (n=4)  Informal consensus (n=10)  Formal consensus (n=6) | 0 (0)  9 (90)  5 (83) | 4 (100)  1 (10)  1 (17) |

* The overall quality AGREE-II score classified the CPG as ‘recommended’ (>60%), ‘recommended with modifications’ (30 to 60%), or ‘not recommended’ (<30%)
