## Supporting information File 2 for "PREOPERATIVE CLINICAL PRACTICE GUIDELINES FOR ELECTIVE SURGERY. METHODOLOGICAL AND QUALITY APPRAISAL STUDY"

**Supplementary Material File 1**

Search Strategies

**Pubmed/MEDLINE**

#1: ((((((((((("guideline"[Publication Type]) OR (Practice Guideline"[Publication Type]")) OR (health planning guidelines[MeSH Terms])) OR (clinical protocols[MeSH Terms])) OR ("consensus development conference"[Publication Type])) OR (Consensus[Mesh])) OR (Standard of Care[Mesh])) OR ("Guideline" [Publication Type])) OR (Guidelines as Topic[Mesh])) OR ("Practice Guideline" [Publication Type])) OR (Health Planning Guidelines[Mesh])) OR (Clinical Protocols[Mesh])

#2: ((((((preoperative) OR (perioperative)) OR (Preprocedural)) OR (surgery)) OR (care, preoperative[MeSH Terms])) OR (period, preoperative[MeSH Terms]))

#3: ((((((cardiac assessment) OR (cardiac evaluation)) OR (cardiac risk)) OR (Cardiovascular Evaluation)) OR (Perioperative Cardiac Risk)) OR (Perioperative Cardiovascular Evaluation))

#4: (fasting[MeSH Terms]) OR (preoperative fasting)

#5:(("preoperative"[All Fields]) OR ("preoperative care"[MeSH Terms])) AND ((("routine"[Title/Abstract]) OR ("test"[Title/Abstract])) OR ("diagnostic tests, routine"[MeSH Major Topic]))

6#: #1 AND #2 AND #3

7#: #1 AND #2 AND #4

8#: #1 AND #2 AND #5

9#: Filters: Publication date from 2010/01/01 to 2022/06/30

**Cochrane Library**

("Guideline" OR "Practice Guideline" OR "health planning guidelines" OR "clinical protocols")

AND ("preoperative" OR "perioperative" OR "preprocedural" OR "surgery" OR "preoperative care")

AND ("cardiac assessment" OR "cardiac evaluation" OR "cardiac risk" OR "Cardiovascular Evaluation" OR "Perioperative Cardiac Risk" OR "Perioperative Cardiovascular Evaluation" OR "fasting" OR "preoperative fasting" OR "preoperative care" OR "routine" OR "test" OR "diagnostic tests")

**EMBASE**

("Guideline" OR "Practice Guideline" OR "health planning guidelines" OR "clinical protocols") AND ("preoperative" OR "perioperative" OR "preprocedural" OR "surgery" OR "preoperative care") AND ("cardiac assessment" OR "cardiac evaluation" OR "cardiac risk" OR "Cardiovascular Evaluation" OR "Perioperative Cardiac Risk" OR "Perioperative Cardiovascular Evaluation" OR "fasting" OR "preoperative fasting" OR "preoperative care" OR "routine" OR "test" OR "diagnostic tests")

**CPG Developers**

1. National Institute for Health and Clinical Excellence ([www.nice.org.uk](http://www.nice.org.uk))
2. Scottish Intercollegiate Guidelines Network ([www.sign.ac.uk](http://www.sign.ac.uk) )
3. New Zealand Guidelines Group ([www.nzgg.org.nz/guidelines](http://www.nzgg.org.nz/guidelines))
4. National Health and Medical Research Council ([www.nhmrc.gov.au](http://www.nhmrc.gov.au))
5. Institute for Clinical Systems Improvement ([www.icsi.org](http://www.icsi.org))
6. Geneva Foundation for Medical Education and Research ([www.gfmer.ch](http://www.gfmer.ch))
7. World Health Organization ([www.who.int](http://www.who.int))
8. UK Department of Health ([www.dh.gov.uk](http://www.dh.gov.uk))
9. National Guidelines Clearinghouse ([www.guidelines.gov](http://www.guidelines.gov))
10. Agency for Healthcare Research and Quality ([www.ahrq.gov/](http://www.ahrq.gov/))
11. ECRI Institute ([www.ecri.org](http://www.ecri.org))
12. American Heart Association ([www.americanheart.org](http://www.americanheart.org))

**Medical Societies**

- World Federation of Societies of Anaesthesiologists (WFSA)
- American Society of Anesthesiologists (ASA)
- European Society of Anaesthesiology (ESA)
- Association of Anaesthetists of Great Britain and Ireland (AAGBI)
- Canadian Anesthesiologists' Society (CAS)
